# Use of the Selective Cytopheretic Device to Support Critically Ill Children Requiring Continuous Renal Replacement Therapy: A Probable Benefit-Risk Assessment

**DOI:** 10.1101/2023.08.22.23294378

**Authors:** Stuart L. Goldstein, Nicholas J. Ollberding, David J. Askenazi, Rajit K. Basu, David T. Selewski, Kelli A. Krallman, Lenar Yessayan, H. David Humes

## Abstract

**Background:** Critically ill children with acute kidney injury (AKI) requiring continuous kidney replacement therapy (CRRT) are at increased risk of death. The selective cytopheretic device (SCD) promotes an immunomodulatory effect when circuit iCa^2+^ is maintained at <0.40 mmol/L with regional citrate anticoagulation (RCA). In a randomized trial of adult patients on CRRT, those treated with the SCD maintaining an iCa^2+^<0.40 mmol/L had improved survival/dialysis independence. We have conducted two multicenter studies to evaluate safety and feasibility of the SCD in critically ill children with AKI receiving CRRT and multiorgan failure. We report the combined efficacy and safety data from these two studies for the first time.

**Methods:** Four pediatric institutions enrolled children ≥10 kg in size with AKI and multiorgan dysfunction (MODS) receiving CRRT as part of standard of care to receive the SCD integrated post CRRT membrane. RCA was used to achieve a circuit iCa^2+^<0.40 mmol/L. We report serious adverse events, patient and CRRT-SCD related process and outcome variables and perform a Bayesian analysis to provide potential attributable benefit-risk assessment of SCD support in this critically ill population using a published matched cohort for the control population.

**Results:** Twenty-two patients (10 females) from the two studies comprise the combined population; 21 received mechanical ventilation, 14 received vasoactive medications, three received extracorporeal membrane oxygenation and 15 had sepsis at the time of CRRT-SCD initiation. Median SCD treatment duration was six days. Fifteen total serious adverse events were recorded, none of which were SCD related. All but one patient survived to the time of SCD discontinuation. Seventeen patients survived 60 days and 16 patients survived to the time ICU discharge. Fourteen patients surviving to ICU discharge had a normal eGFR and no patient was dialysis dependent at 60 days after CRRT-SCD initiation. Bayesian analyses revealed a 98-99% probable benefit of addition of SCD support.

**Conclusion:** These data suggest SCD therapy is feasible, safe and demonstrates probably benefit for children who require CRRT for AKI in the setting of MODS.

## Introduction

Acute kidney injury (AKI) is a significant complication in critically ill children, as it results in increased morbidity and mortality.^1–4^ AKI develops predominantly due to the injury/necrosis of renal proximal tubule cells, often concurrently with sepsis. Part of the disease process in patients with AKI is often the development of a systemic inflammatory response syndrome (SIRS), resulting in cardiovascular collapse, ischemic damage to vital organs and multi-organ dysfunction (MODS).^5, 6^

Despite advancements in renal replacement therapy (RRT), mortality rates in children with AKI **and** MODS requiring continuous RRT (CRRT) continues to approach 50%.^7–9^ Children who survive an AKI episode are at increased risk of chronic kidney disease (CKD).^10–12^ Since activated leukocytes are central to the pathogenesis/progression of septic shock and other clinical inflammatory disorders new therapeutic approaches are being considered to limit the deleterious clinical effects of activated leukocytes.^13^

The selective cytopheretic device (SCD, SeaStar Medical, Inc., Denver, Colorado) immunomodulates activated circulating leukocytes and provides a new therapeutic approach to SIRS and AKI. We have previously published SCD results from: 1) a prospective US Food and Drug Administration (FDA) funded, multi-center safety and feasibility study^14^ in children ≥20 kg 2) a case-report^15^ of a 21-month-old with EBV driven HLH and 3) a case series of three children with Shiga-toxin producing *Escherichia coli* associated hemolytic uremic syndrome (STEC-HUS).^16^ We now compile data from all these patients and others into a single report to summarize the entire pediatric patient experience with the SCD. Our aim is to support the premise that SCD treatment is safe, with probable benefit in this critically ill pediatric population.

## Methods

We conducted two prospective studies (SCD PED-01 and SCD PED-02) at four US centers (Cincinnati Children’s Hospital Medical Center, University of Michigan/ CS Mott Children’s Hospital, University of Alabama at Birmingham/ Children’s of Alabama, and Emory University/Children’s Healthcare of Atlanta at Egleston). Children of ≥20 kg and up to 22 years of age (SCD PED-01), and ≥10-20 kg (SCD PED-02) admitted to an intensive care unit (ICU) were screened for eligibility. Subjects who had AKI, as defined by the Kidney Disease Improving Global Outcomes (KDIGO) criteria^17^ and MODS receiving CRRT as part of the standard of clinical care were eligible to be enrolled. MODS was defined as respiratory disease requiring invasive mechanical ventilation and/or cardiovascular compromise requiring the provision of a continuous infusion of an inotropic/vasoactive medication. Pediatric Risk of Mortality score (PRISM III) at ICU admission was used to assess severity of patient illness.^18^ The Institutional Review Board at each center approved the study prior to patient enrollment at their site. Written informed consent was obtained from the subject’s parents/person with medical decision-making power prior to subject enrollment.

The studies received an IDE (G120174) from the US FDA and were registered at www.clinicaltrials.gov (SCD PED-01: NCT02820350, SCD PED-02: NCT04869787) prior to each study’s commencement. SCD PED-01 was funded mostly by an Office of Orphan Products Development (OOPD) grant (R01FD005092) from the US FDA with a small subsequent grant from SeaStar Medical Inc. to complete the final two subjects’ enrollment. SCD PED-02 was funded by the Frankel Innovation Initiative at the University of Michigan. As these are primarily safety studies (adverse events (AE) and serious adverse events (SAE)), results and progress were reviewed by an external Data Safety Monitoring Board (DSMB) at least annually and within 5 days for any SAE.

### CRRT-SCD protocol

The pediatric SCD protocol has been published extensively elsewhere.^14^ All centers provided CRRT with RCA as part of their local standard of clinical care with the following study required constraints: 1) small solute clearance was required to be at least 2000 mL/hour/1.73m^2^ of patient Body Surface Area (BSA), 2) RCA was provided with a well-described protocol^19–21^ using Anti-Coagulant Dextrose-A (ACD-A™, Baxter Healthcare) to maintain the CRRT circuit iCa^2+^ <0.40 mmol/L for at least 90% of the time a subject was receiving CRRT-SCD 3) CRRT circuit and subject systemic iCa^2+^ were measured at least every six hours and 4) only polysulfone based CRRT membranes could be used (i.e., no AN-69 membranes).

### Analyses

The primary outcome of these studies was to evaluate the safety of the SCD in children weighing over 10 kg to support and application for an HDE. Subject demographic data and CRRT/SCD data are aggregated and reported as median and interquartile range (IQR). AEs and SAEs were reported by the site investigators based on pre-determined criteria. The site investigator determined if any AE was related to the device. An AE, whether considered study-treatment related or not, which fit any of the criteria below, was considered a SAE:

- Resulted in death
- Was life-threatening (meaning that the patient was at risk of death at the time of the event; this does not refer to an event which might have caused death if it had occurred in a more severe form)
- Required in-patient hospitalization or prolongs the existing hospitalization
- Was a persistent disability/incapacity
- Was considered an important medical event by the investigator (e.g., surgery, return to ICU, emergency procedures)

The secondary outcomes were: patient survival to ICU discharge or to 60 days after CRRT (CRRT-SCD) initiation and estimated glomerular filtration rate (eGFR) at 28 and 60 days after CRRT-SCD initiation. The serum creatinine based bedside Schwartz equation^22^ was used to calculate eGFR:

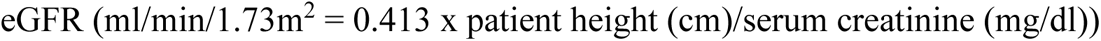

To assess for probable benefit, patient survival to ICU discharge or 60 days in the combined cohort was compared to a subset from the Prospective Pediatric Continuous Renal Replacement Therapy (ppCRRT) Registry^7^ with patients ≥10 kg who received either invasive mechanical ventilation or a vasoactive medication at the time of CRRT initiation. The ppCRRT Registry enrolled a total of 370 patients; the comparison subset of 210 patients included all patients who met the matching criteria (i.e., there were no missing data in the ppCRRT to lead to a selection bias). Initial comparison was performed using chi-square analysis with Fisher Exact test. To validate this crude analysis, Bayesian logistic regression, as implemented by the brms package (version 2.17.0), was used to estimate the probability that the log odds of survival to 60 days in the SCD cohort exceeded that observed in the ppCRRT cohort. Models were fit assuming default flat (e.g., uniform) priors and a logit link function to model the Bernoulli distributed responses. A total of 8,000 iterations were carried out across four chains with a burn-in of 500 iterations per chain using the STAN No-U-Turn Sampler and model convergence assessed via trace plots and r-hat values. Posterior samples were transformed to the probability scale using the inverse logit transform and used to compute the predicted probability of surviving to 60 days for each cohort, the probability of treatment benefit (e.g., log odds SCD > ppCRRT), and the predicted risk difference. All analyses were carried out using the R software environment for statistical computing and graphics (version 4.1.1) and rstan (version 2.21.3).

## Results

The pre-SCD initiation, SCD procedure and patient outcome data are shown in aggregate for the 22 patients and divided between the two studies in **Table 1**. Aggregate patient outcomes from CRRT-SCD initiation to Day 60 post-initiation are depicted in **Figure 1**.

**Figure 1.**
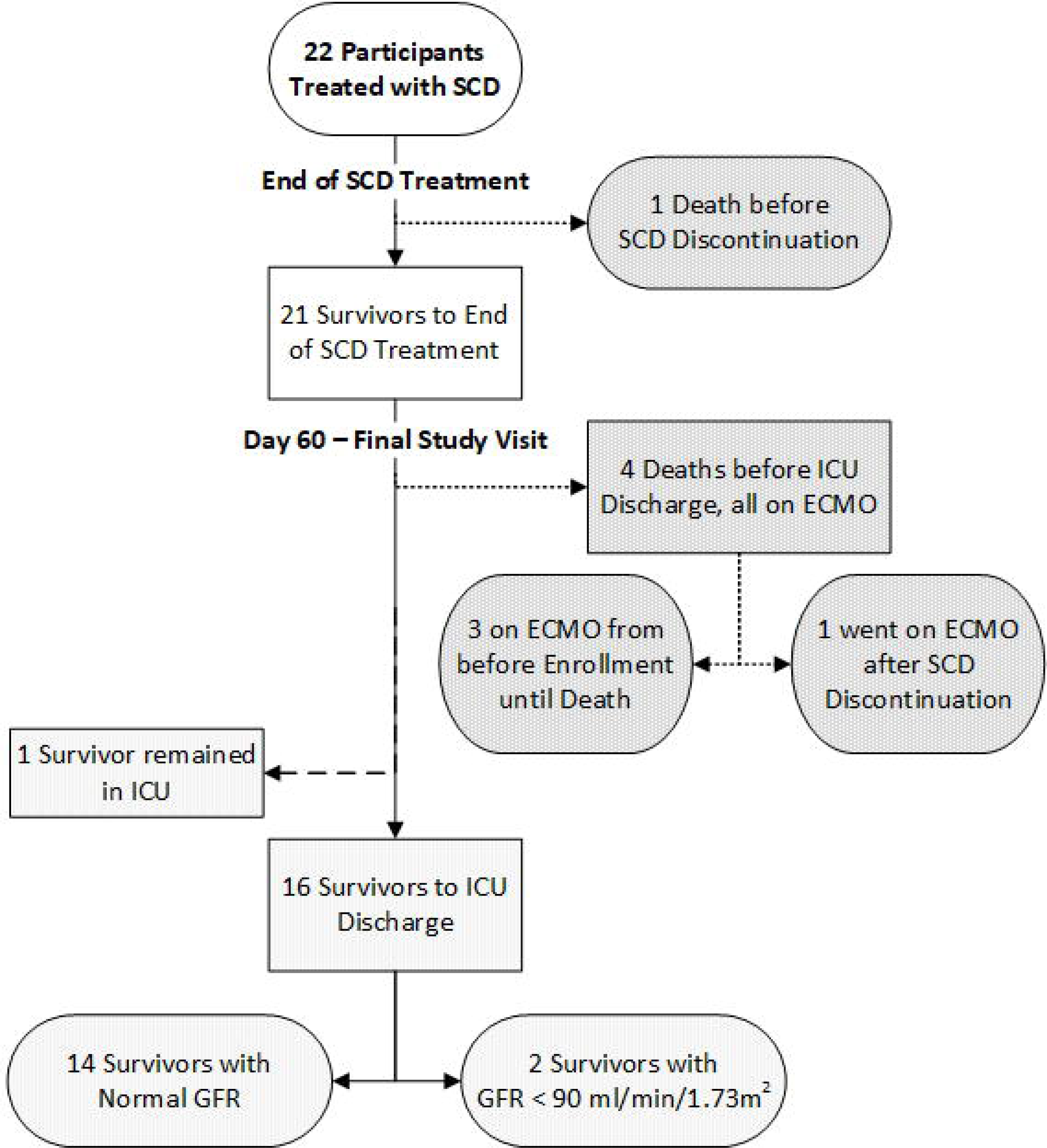
Flow and outcomes for subjects enrolled in the SCD-PED -01 and SCD- PED-02 studies.

**Table 1.**

| Patient Demographics and Pre-SCD Characteristics |  |  |  |
| --- | --- | --- | --- |
|  | Combined<br>(n=22) | SCD-PED-01<br>(n=16) | SCD-PED-02<br>(n=6) |
| Patient Sex (F/M) | 12/10 | 8/8 | 2/4 |
| Patient Age (years) | 9.5 [ 4.3, 15.8] | 12.1 [7.8, 16.8] | 2.1 [1.7, 3.5] |
| Patient Weight (kg) | 30.3 [16.5, 62.1] | 53.3 [27.1, 69.5] | 13.4 [12, 14.1] |
| PRISM III at ICU admission | 9.5 [7, 14] | 10.5 [6.5, 14.5] | 8.5 [7, 13] |
| Mechanical Ventilation at SCD Start | 21 | 15 | 6 |
| Vasoactive Medication at SCD Start | 14 | 9 | 5 |
| ECMO at SCD Initiation | 3 | 2 | 1 |
| ECMO at SCD Initiation or After | 4 | 3 | 1 |
| Sepsis at SCD Initiation | 15 | 10 | 5 |
| Fluid Accumulation to CRRT Start (%) | 7.5 [3.0, 14.5] | 5.1 [2.8, 9.7] | 23.6 [12.5, 31.6] |
| ICU Days to CRRT Start | 3.5 [1, 5] | 2 [1, 4.5] | 4.5 [4, 8] |
| CRRT-SCD Procedure Characteristics |  |  |  |
|  | Combined<br>(n=22) | SCD-PED-01<br>(n=16) | SCD-PED-02<br>(n=6) |
| CRRT Days before SCD | 4 [0, 4.8] | 4 [0, 4.5] | 7.8 [0, 25.8] |
| Days on CRRT-SCD | 6 [3, 7] | 6 [4, 7]<sup>a</sup> | 5 [1, 9]<sup>b</sup> |
| CRRT Days after SCD | 0 [0, 4] | 0 [0, 4.5] | 0 [0,0] |
| SAEs Total | 15 | 9 | 6 |
| CRRT-SCD Patient Outcomes |  |  |  |
|  | Combined | SCD-PED-01 | SCD-PED-02 |
| Survival to SCD discontinuation | 21/22 | 15/16 | 6/6 |
| Survival to CRRT Discontinuation | 21/22 | 12/16 | 5/6 |
| Survival to ICU Discharge | 16/21 | 12/16 | 4/5 |
| Survival to 60 days<sup>c</sup> | 17/22 | 12/16 | 5/6 |
| Survival to ICU Discharge or 60 days in non-ECMO patients<sup>d</sup> | 17/18 | 11/12 | 6/6 |
| Day 28 estimated GFR (ml/min/1.73m<sup>2</sup>) | 90 [45, 140] | 80 [37, 109] | 104 [80, 240] |
| Day 60 Estimated GFR (ml/min/1.73m<sup>2</sup>) | 105 [97, 119] | 105 [99, 114] | 111 [95, 180] |

- a. SCD-PED-01 protocol had maximum SCD treatment of seven days. - b. SCD-PED-02 protocol had maximum SCD treatment of ten days. - c. One patient alive and still in ICU at 60 days - d. All four patients receiving ECMO during their ICU course died.

All but one patient survived until the time SCD and CRRT discontinuation. The one deceased patient (reported previously) in SCD PED-01 died seven hours after CRRT-SCD initiation and was found to have extensive invasive viral myocarditis on biopsy. The DSMB and FDA each reviewed the case and assessed the death to not be related to the SCD. Sixteen patients survived until the time of ICU discharge; four of the five non-survivors received ECMO support at some point during their ICU course (three while on CRRT-SCD, one after CRRT-SCD completion). One patient was still in ICU on Day 60, 14/16 ICU survivors had an eGFR >90 ml/min/1.73m^2^ and no patient was dialysis dependent on Day 60 after CRRT-SCD initiation.

While no claims can be made regarding efficacy with such a small patient sample size, an HDE study must show a device safe with probable benefit for marketing clearance. To assess for probable benefit, we compared the combined cohort to a size matched cohort from the (ppCRRT) registry (**Table 2**). No differences were observed between the two cohorts with respect to patient characteristics or severity of illness at the time of CRRT initiation. One hundred fifteen of 210 patients from the matched ppCRRT cohort and 17 of the 22 patient SCD combined cohort survived to the time of ICU discharge or 60 days (55%, 95%CI 48-62% *vs* 77%, 95%CI 55-92%, p=0.04). To validate this crude comparison, we undertook a Bayesian analysis comparing the two cohorts. The distribution of the predicted probabilities for survival to the time of ICU discharge or 60 days obtained from the posterior samples is shown in **Figure 2A** for each cohort separately. In 98% of the posterior samples, the log odds of surviving to the time of ICU discharge or 60 days was greater for the SCD when compared to the ppCRRT cohort (**Figure 2B**); highlighting the probable benefit in this cohort of patients. The predicted risk difference was 22.4% (95% credible interval 2.5%; 38.5%) for the SCD versus ppCRRT cohort (**Figure 2C**). A similar estimate of the probable benefit (99%) was obtained by directly estimating the percentage of times the mean of a binomially distributed sample of n=22 observations with a probability of 17/22 exceeded that of one drawn from a sample of n=210 observations and probability of 115/210 when taken over 1 x 10^6^ samples.

**Figure 2.**
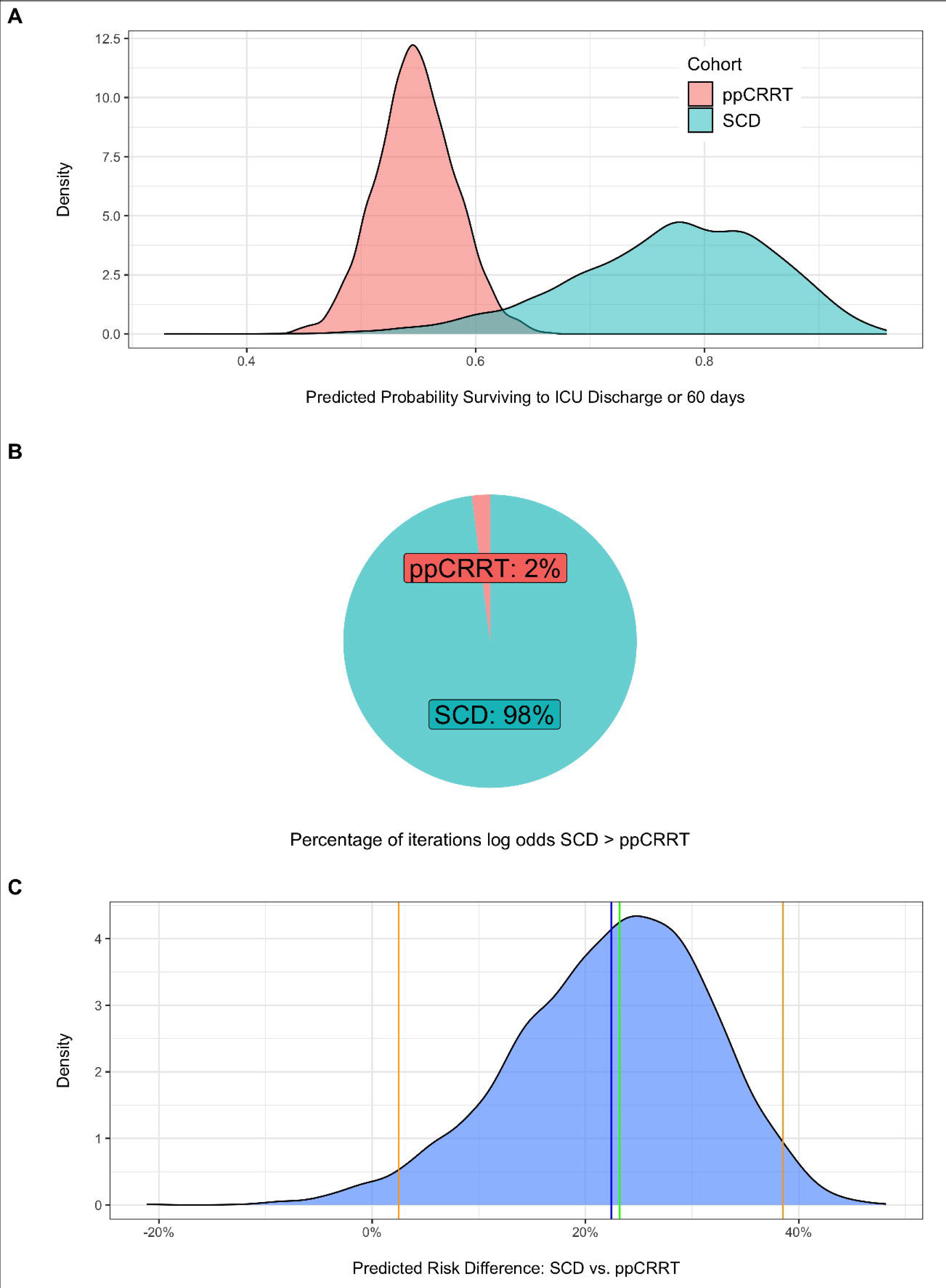
Predicted probabilities of survival to ICU discharge or 60 days. 2A). Posterior predicted probabilities of survival to ICU discharge or 60 days for the SCD (blue) and ppCRRT (red) cohorts. 2B.) The percentage of posterior samples in which the log odds of surviving to ICU discharge or 60 days was greater for the SCD cohort than the ppCRRT Registry cohort. 2C). Predicted risk difference computed from the posterior samples. Mean difference shown in blue. Median difference shown in green. 95% credible interval shown in orange.

**Table 2.** SCD and ppCRRT Population Comparisons.

|  | **ppCRRT**<br>**(n=210)** | **CRRT-SCD**<br>**(n=22)** | **p-value** |
| --- | --- | --- | --- |
| **Patient Sex (F)** | 94 (44.7%) | 10 (45.4%) | 0.91 |
| **Patient Age (years)** | 11.3 [5.0, 16.4] | 9.5 [ 4.3, 15.8] | 0.50 |
| **Patient Weight (kg)** | 36.9 [19, 60] | 30.3 [16.5, 62.1] | 0.86 |
| **PRISM II at ICU admission** | 12 [8, 18] | 11.5 [5, 15] | 0.11 |
| **Mechanical Ventilation at CRRT Start** | 183 (87.1%) | 21 (95.4%) | 0.21 |
| **Pressors at CRRT Start** | 160 (76.1%) | 15 (68.1%) | 0.48 |
| **CRRT Duration (Days)** | 7 [3, 13] | 6.5 [5, 11.5] | 0.88 |
| **Survival to ICU Discharge or Day 60** | 115 (54.8%) | 17 (77.3%) | **0.04** |

## Discussion

We report aggregate data from two pediatric studies using the SCD to support critically ill children with AKI, MODS who received CRRT as part of their standard of care. Our data suggest that SCD therapy was safe and feasible in this cohort, as no device related SAEs were observed, and that there was a high likelihood of benefit of SCD therapy in this critically ill pediatric population.

The precise mechanism of action of the SCD is becoming better understood and appears to be an immunomodulatory process which inhibits leukocyte activation, a critical component of the SIRS leading to MODS. The modulation of the dysregulated inflammatory state also allows recovery of renal function in AKI and other associated organ failures. The cartridge acts as a selective cytopheretic device in the presence of citrate anticoagulant to bind and immunomodulate potentially damaging circulating leukocytes. This perspective is based upon evolving data from *in vitro* bench studies, preclinical animal models, and human clinical trials utilizing measurements of inflammatory biomarkers and leukocyte cell sorting, cytometric analysis. More complete discussions of the possible mechanisms can be found elsewhere.^23–25^

Given the limited sample size of 22 subjects, we can make no claims about the efficacy of the SCD on patient related outcomes. However, the 77% ICU survival rate observed compares favorably to published CRRT studies in critically ill children with MODS that show a survival rate of about 50%. The largest multicenter study from the ppCRRT Registry who received mechanical ventilation or a vasoactive agent, which were identical inclusion of SCD-PED-01 and SCD-PED-02 showed an ICU survival rate of 51.7%.^9^ Notably, ECMO provision was an exclusion in the ppCRRT, so a potentially more relevant comparison is a combined SCD ICU survival rate of 16/17 (94.1% excluding the four patients receiving ECMO in the CRRT- SCD cohort) *vs.* 51.7% in the ppCRRT cohort. In addition, our analyses comparing survival to the time of ICU discharge or 60 days for patients using the SCD versus those receiving mechanical ventilation or a vasoactive agent in the ppCRRT Registry suggests a potential probability of treatment benefit as high as 98% for those receiving SCD. Thus, these results support the likelihood of probable benefit for this device when compared to standard treatment, which is a requirement component for Humanitarian Device Exemption clearance by the US FDA. Currently, the FDA is reviewing the HDE application for SCD-PED-01,^14^ where similar analyses for that 16-patient cohort demonstrated an 88-92% likelihood of probable benefit in comparison to a matched ppCRRT cohort.

In summary, we demonstrated a high level of safety of SCD therapy and probable efficacy in children ≥10 kg with AKI and MODS receiving CRRT standard of care and suggest a favorable benefit to risk ratio in this critically ill pediatric population. The US FDA Office or Orphan Products Development granted SeaStar Medical, Inc. a Humanitarian Use Designation for the SCD under the study indicated use. Further studies will be required to establish safety in smaller children, feasibility of integration with other CRRT platforms, and to demonstrate improved patient outcomes compared to current supportive therapy which is associated with a high rate of CKD in survivors.

## Data Availability

All data produced in the present study are available upon reasonable request to the authors

